# The importance of vaccinated individuals to population-level evolution of pathogens

**DOI:** 10.1101/2022.11.29.22282822

**Authors:** Maria A. Gutierrez, Julia R. Gog

## Abstract

Virus evolution shapes the epidemiological patterns of infectious disease, particularly via evasion of population immunity. At the individual level, host immunity itself may drive viral evolution towards antigenic escape. Using compartmental SIR-style models with imperfect vaccination, we allow the probability of immune escape to differ in vaccinated and unvaccinated hosts. As the relative contribution to selection in these different hosts varies, the overall effect of vaccination on the antigenic escape pressure at the population level changes.

We find that this relative contribution to escape is important for understanding the effects of vaccination on the escape pressure and we draw out some fairly general patterns. If vaccinated hosts do not contribute much more than unvaccinated hosts to the escape pressure, then increasing vaccination always reduces the overall escape pressure. In contrast, if vaccinated hosts contribute significantly more than unvaccinated hosts to the population level escape pressure, then the escape pressure is maximised for intermediate vaccination levels. Past studies find only that the escape pressure is maximal for intermediate levels with fixed extreme assumptions about this relative contribution. Here we show that this result does not hold across the range of plausible assumptions for the relative contribution to escape from vaccinated and unvaccinated hosts.

We also find that these results depend on the vaccine efficacy against transmission, particularly through the partial protection against infection. This work highlights the potential value of understanding better how the contribution to antigenic escape pressure depends on individual host immunity.

## 1 Introduction

Vaccines against infectious pathogens reduce incidence and deaths, although there is a risk that they could favour new antigenic variants [1]. For example, since population immunity against circulating SARS-CoV-2 variants controls virus spread [2], the long-term dynamics of the COVID-19 pandemic depend on the evolutionary trajectory of the virus [1]. Here we investigate how vaccination changes the appearance probability of strains capable of evading host immunity, relative to the baseline of no vaccination. Antigenic escape is complex, with strains generated and selected on various scales [3]. We focus exclusively on the *generation* of escape strains [4], [5], rather than on their establishment after they first appear[6], [7]. Our work builds on other mathematical studies of COVID-19 immune escape risk [4], [5], [8]. We pay particular attention to selection in vaccinated hosts, relative to the unvaccinated.

Evolution of antigenic traits in SARS-CoV-2 (severe acute respiratory syndrome coronavirus 2) [9] generates new variants [10]. Some of these variants (B.1.1.7, *alpha*; B.617.2, *delta*) appear to have a transmission advantage over their precursors [11]. B.1.1.529 —*omicron*—significantly escapes existing immunity [12], but variants with immune escape, at least to some extent, date back to 2020, before the roll-out of vaccines (B.1351, *beta*; B.617.2, *delta*) [11]. Therefore it is important to understand how vaccination impacts antigenic drift. Vaccines may affect the evolution of other pathogen traits, such as virulence [13], but we focus on immune escape.

To the best of our knowledge, there is not yet evidence to assess how COVID-19 vaccines impact virus genomic diversity in an individual host [14]. Some COVID-19 models assume that escape occurs only or primarily in hosts with previous immunity [4], [5]. On the contrary, others assume that vaccination entirely prevents infection [8], and thus escape can only occur in unvaccinated hosts.

Models for pathogens other than SARS-CoV-2 often assume that viruses mutate at the same rate in all hosts [7], [15], [16], [17]. However, in human Influenza A, host immunity can select for antigenic mutations [18]. This effect may be important [19] for more ambitious vaccination campaigns, such as a universal influenza vaccine [20].

In this paper, we follow the minimal approach of others (for example, [4]) to define an escape pressure function proportional to the number of infections. Given that we do not generally know the relative selective pressures exerted by infections in vaccinated hosts compared to the unvaccinated, we explore a range of values for this contribution to escape. We define a parameter (Section 2.2) that measures this relative contribution to escape. We use this parameter to express the escape pressure in terms of the infections in vaccinated and unvaccinated hosts. We combine this with deterministic compartmental models (Section 2.1) for vaccination that provides lifelong imperfect immunity. We explore two scenarios: a single wave and endemic disease. We calculate the resultant escape pressure in each of these contexts and describe how it changes as the vaccination coverage varies. We find the tradeoff between selection and the vaccine efficacies (VEs) in the escape pressure of a fully-vaccinated population and the vaccination coverage that maximises the escape pressure (Section 3).

We find that the relative selection strength from infections in vaccinated and unvaccinated hosts shapes the escape pressure. In particular, we find two different qualitative behaviours for how the escape pressure depends on the vaccination coverage. These two behaviours are separated by a threshold value of this relative contribution to escape, in which vaccinated hosts contribute individually somewhat more to escape than the unvaccinated. If the relative contribution to escape is below this threshold, vaccination always reduces antigenic escape, as in [15]. However, above this threshold, intermediate vaccination levels are the most likely to generate escape strains, as in [4]. We also find that the susceptibility reduction provided by the vaccines has a large effect on the escape pressure. The reduction in infectiousness is less important, but still lowers escape by reducing prevalence.

## 2 Methods

### 2.1 Epidemic models

We use systems of ordinary differential equations (ODEs) to model disease transmission. We assume that the population is well-mixed and a single-strain infectious disease circulates unaffected by any escape strain it may have generated. We consider two epidemiological scenarios. In scenario (a) the outbreak is transient, which can be interpreted as a single epidemic wave. In scenario (b) the system is modified so that it reaches a non-zero equilibrium, corresponding to endemic disease.

We separate individuals by vaccination status and assume that vaccine immunity does not wane. We scale time so that the recovery rate is 1. In our models, the vaccines are not assumed to be perfect: vaccinated individuals can become infected, albeit at a lower rate, contributing to the escape pressure. We split the (imperfect) transmission blocking from vaccines into two components, *θ*_*S*_ and *θ*_*I*_, as defined in [4]. *θ*_*S*_ is the susceptibility reduction (VE against infection): *θ*_*S*_ = 0 corresponds to perfect protection, while *θ*_*S*_ = 1 corresponds to no protection. Similarly, *θ*_*I*_ is the infectivity reduction (VE against transmission), if an infection occurs. For mathematical convenience, the reduction in susceptibility from vaccination is polarised [16]: some vaccinated individuals are entirely immune to the infection, while the rest are as susceptible as the unvaccinated. Appendix A.2 shows that the same qualitative results hold with an alternative formulation for immunity.

In both scenarios, *S*_*U*_ and *S*_*V*_ are the vaccinated and unvaccinated proportions of the population susceptible to the disease; *I*_*U*_ and *I*_*V*_, the vaccinated or unvaccinated proportions that are infected; *R*_*U*_ and *R*_*V*_, the vaccinated or unvaccinated recovered proportions. *R*_0_ is the basic reproduction number and *c* is the proportion of the population who are vaccinated.

#### Scenario (a): transient epidemic

In the transient scenario, recovered hosts have full permanent immunity and the system is

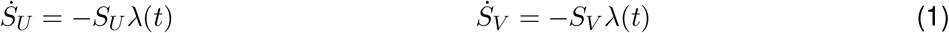

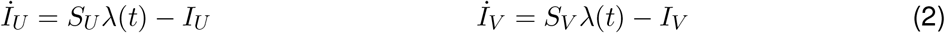

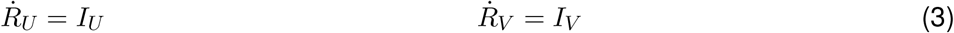

with *λ*(*t*) = *R*_0_(*I*_*U*_ + *θ*_*I*_*I*_*V*_), the force of infection. The initial conditions are *S*_*U*_ = 1 −*c, S*_*V*_ = *cθ*_*S*_, *R*_*U*_ = 0 = *R*_*V*_, and an infinitesimal number of infected individuals. The remaining *c*(1 −*θ*_*S*_) proportion of the population has full immunity from vaccination, so they do not appear in the compartments of the system. Appendix A.1 shows that for polarised vaccine immunity —and non-assortiative mixing as here—, the original proportion between vaccinated and unvaccinated susceptibles is maintained and extended to the other compartments:

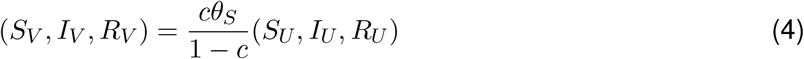

This relation (4) reduces the system to:

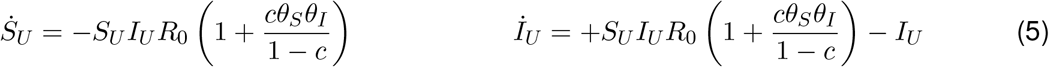

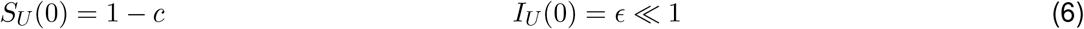

where *ϵ* accounts for the initial cases. (5)-(6) describe a standard SIR model, up to rescaling the effective reproduction number and the population size. The initial effective reproduction number is *R*_*e*_ = *R*_0_[1 − *c* + *cθ*_*S*_*θ*_*I*_]. If the vaccination coverage is above 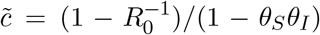, herd immunity prevents an outbreak and prevalence decreases exponentially. From (5)-(6), if 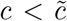, the cumulative infections in unvaccinated hosts are 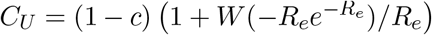, where *W* is the Lambert-W function [21] (see Appendix A.1). Using (4), the cumulative breakthrough infections are 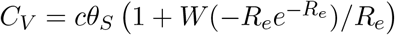.

#### Scenario (b): endemic disease

To achieve an endemic scenario, we consider two possible extensions of the SIR model (1)-(3) that replace susceptibles. If starting from the same initial conditions as in the transient scenario, these extensions reach an endemic state if *R*_*e*_ *>* 1. In both cases, we find the proportions 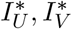 of infected individuals at the endemic equilibrium. Both models lead the same expressions for the prevalences 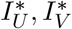, up to an overall constant of proportionality 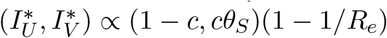.

In the first extension, we assume that immunity from infections wanes at a constant rate *ω*, instead of being permanent. We still assume that vaccine immunity is lifelong. The modified ODEs are

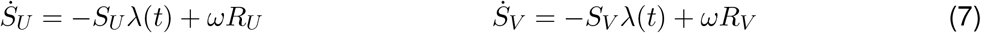

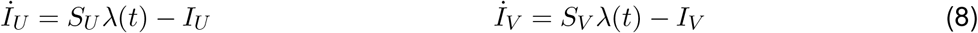

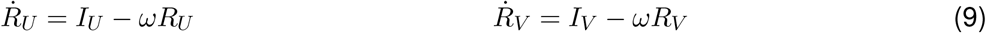

with *λ* as before. We use (4), which still holds (see Appendix A.1), to find the endemic state (setting to zero the time-derivatives). For *R*_*e*_ *>* 1, there is a stable equilibrium state with 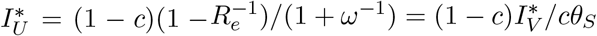.

In the second extension, we include births and deaths, at the same homogeneous per capita rate *μ*, so that the population size is conserved. A proportion *c* of the population is vaccinated at birth. We assume that neither infection nor immunity are maternally transmitted. Therefore, the births appear in 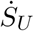 and 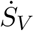 as *μ*(1 − *c*) and *μcθ*_*S*_. The number of individuals *V* with full vaccine immunity obeys 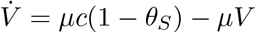 with *V* (0) = *c*(1 − *θ*_*S*_), so *V* (*t*) = *c*(1 − *θ*_*S*_) at all times. The new ODEs are

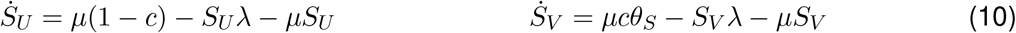

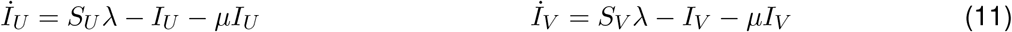

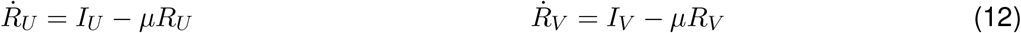

with *λ* as before. Condition (4) still holds (see Appendix A.1). For *R*_*e*_ *>* 1 + *μ*, there is an endemic state 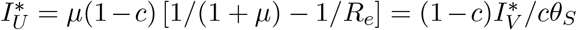. For *μ* ≪ 1, which corresponds to an infectious period much shorter than the host’s life expectancy, 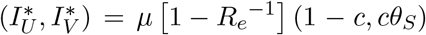 to leading order in *μ*.

### 2.2 Escape pressure

We take a simple approach building on [4] to study the generation of strains at the population-level. We assume that the escape pressure *P* depends linearly on the number of cases at that time, as

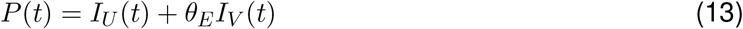

Underlying this factor is the assumption that each infection contributes very slightly to escape potential. We weight the (breakthrough) infections in vaccinated hosts *I*_*V*_, relative to the infections in naïve hosts *I*_*U*_, by a factor *θ*_*E*_. Only the relative value of *P* is important (see equations (16), (17)), so we only need the relative weighting *θ*_*E*_ amongst infection in the vaccinated and unvaccinated rather than their absolute contribute to escape. *θ*_*E*_ is comparable to the vaccine efficacies *θ*_*S*_, *θ*_*I*_ of Section 2.1, in that it is a factor that reflects a change in a single host induced by vaccination. For example, if *θ*_*E*_ *>* 1, vaccinated hosts infected with the resident strain would be more likely to pass on escape strains than the unvaccinated. Lower viral loads (*θ*_*I*_ < 1) and shorter infections in the vaccinated [22] could lead to fewer mutations [23]. However, escape strains may avoid the vaccine-induced immunity that the original strain faces [24], and may be selected within a host enough to be transmitted to others [23]. There is no prior reason to exclude any value *θ*_*E*_ ≥ 0, so we explore the full range.

The escape pressure as in equation (13) is time-dependent. *P*(*t*) scales roughly with the total number of cases. For the models of Section 2.1 with polarised immunity, the number of infected individuals in the unvaccinated and vaccinated compartments, *I*_*U*_(*t*), *I*_*V*_ (*t*) are proportional to each other (equation (4)). Therefore, for fixed *θ*_*E*_ and vaccination coverage, *P*(*t*) is proportional to the prevalence. This scaling fails if equation (4) does not hold (for example in the model with leaky immunity of Appendix A.2). In general, the escape pressure as a function of time is a linear superposition of the curves for infections in vaccinated and unvaccinated hosts. Nonetheless, our main interest is how the escape pressure depends on the vaccination coverage *c* and *θ*_*E*_. Hence, we need a reasonable way to eliminate the time-dependency of *P*(*t*). For the transient scenario, we consider the cumulative escape pressure throughout the full wave 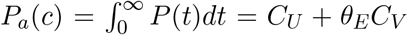. For the endemic scenario, we study the asymptotic value of *P* at the endemic equilibrium 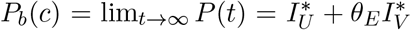. With the expressions for 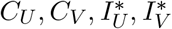 from Section 2.1,

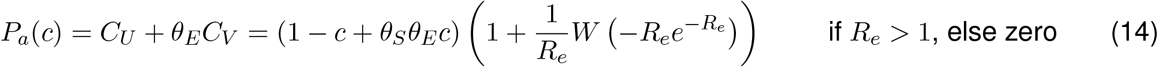

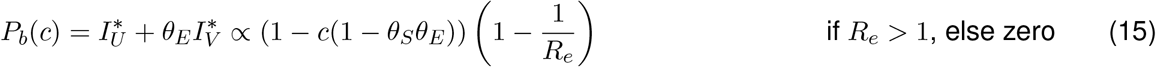

To study the differential effect of the vaccination coverage on the escape pressure, we normalise the expressions (14) and (15) by their value at *c* = 0 (in the absence of vaccination):

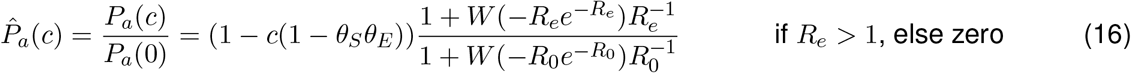

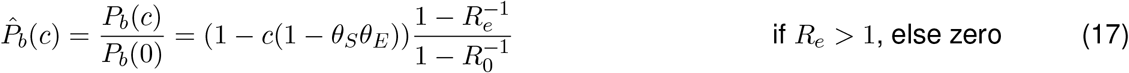

If 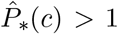 (where ∗ is *a* or *b*), the vaccination coverage *c* increases the escape pressure from no vaccination (in scenario (*a*) or (*b*), respectively). Expressions (16), (17) are similar, but 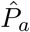 and 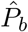 should not be compared between each other. They correspond to distinct epidemiological scenarios and have different derivations from the time-dependent escape pressure *P*(*t*) of equation (13).

## 3 Results

First, we focus on the effects of *θ*_*E*_ on the escape pressure in a fully vaccinated population. We vary *θ*_*S*_, *θ*_*I*_ and *θ*_*E*_, and fix the vaccination coverage at *c* = 1. If vaccination does not prevent the epidemic, i.e., *R*_0_*θ*_*S*_*θ*_*I*_ < 1:

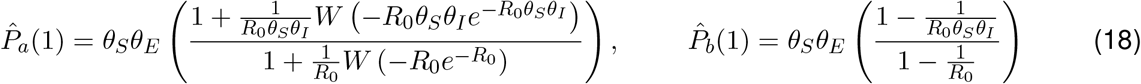

and 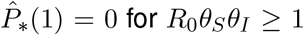. Both expressions in (18) depend only on *R*_*0*_, *θ*_*S*_*θ*_*I*_ and *θ*_*S*_ *θ*_*E*_, but this dependence changes for “leaky” immunity (see Appendix A.2). Figures 1a, S1a show how the escape pressure in a fully vaccinated population 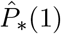 depends on the vaccine parameters *θ*_*S*_, *θ*_*I*_ and *θ*_*E*_. As expected, it is unchanged relative to no vaccination 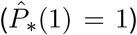 when *θ*_*E*_ = 1 = *θ*_*S*_ = *θ*_*I*_, because then vaccines have no effect in our model. For a fixed VE, the escape pressure increases with *θ*_*S*_*θ*_*E*_, because escape becomes more likely in the vaccinated. Similarly, for fixed *θ*_*S*_*θ*_*E*_, the escape pressure decreases as the vaccines become more effective, because fewer infections reduce the opportunities for escape mutations. Therefore, the escape pressure in a fully-vaccinated population is less than in an unvaccinated population 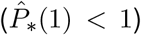 if the population is close enough to herd-immunity. As a consequence, *θ*_*S*_ *θ*_*E*_ < 1 is not necessary for 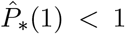. It is sufficient, because if *θ*_*S*_ *θ*_*E*_ < 1, vaccination blocks the expected escape so 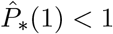 unsurprisingly. Our results shows that vaccines can reduce the total escape pressure even if *θ*_*S*_*θ*_*E*_ *>* 1.

**Figure 1:**
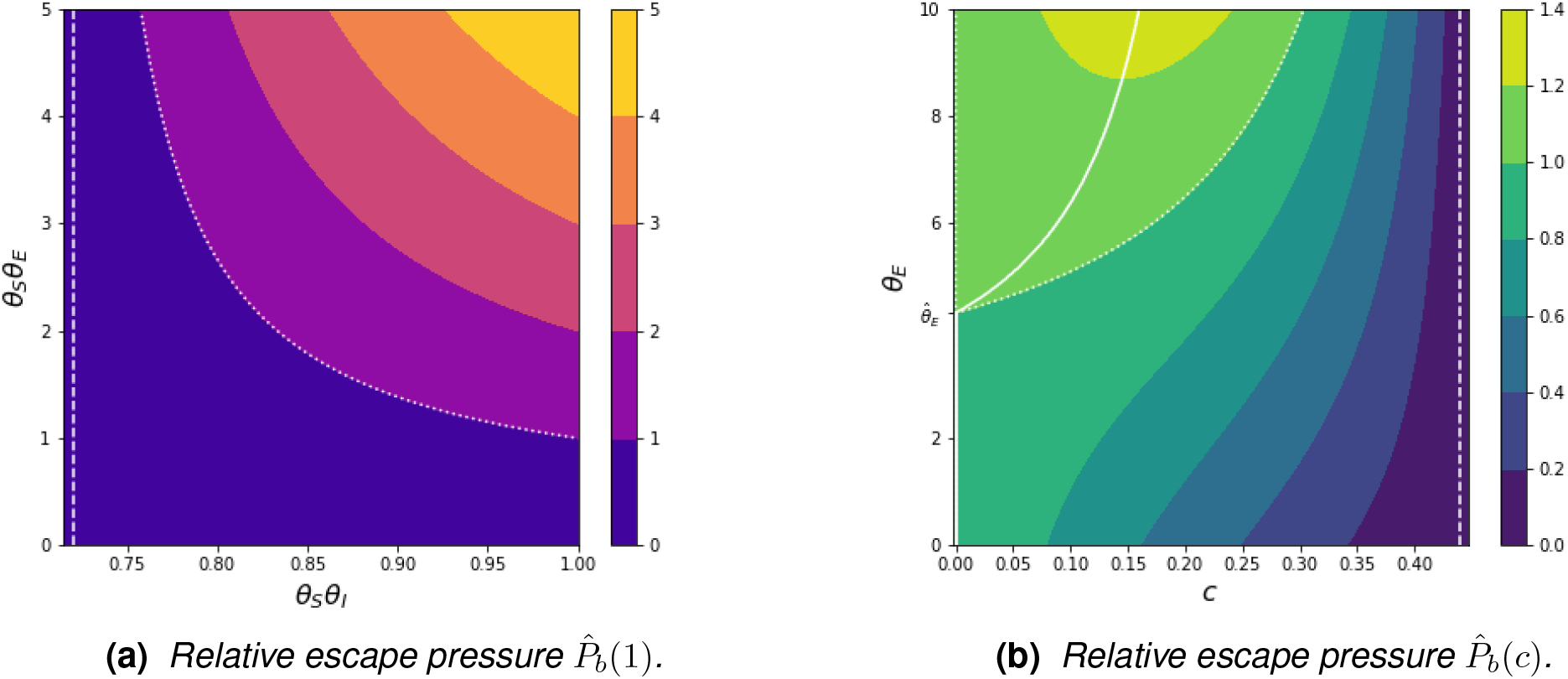
(a): Escape pressure 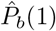 in a fully vaccinated population relative to an unvaccinated population. White dotted curve 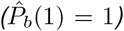: vaccine parameters that give the same escape as in an unvaccinated population. White dashed line: VE that prevents an epidemic in a fully vaccinated population (the horizontal axis is scaled to above this value). R_0_ = 1.4. (b): Escape pressure 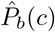 for variable vaccination coverage c and relative contribution to escape θ_E_ from the vaccinated, relative to an unvaccinated population. White solid curve: the vaccination coverage, c_m_, at which the escape pressure peaks, as a function of θ_E_. White dotted curve 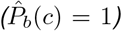: combination of c and θ_E_ that give the same escape in a population as in an unvaccinated population. Dashed line: vaccination coverage for herd-immunity threshold (the horizontal axis only shows values of c below this). θ_S_, θ_I_ = 0.6, R_0_ = 1.4.

**Table 1:** Parameters and functions from our model that appear in the results section. For the input vaccine and epidemiological parameters, we specify their ranges of feasible values. The realised values of the output parameters (bottom block of the table) differ between scenarios.

| category | item | description |
| --- | --- | --- |
| vaccine parameters | $\theta_S \in [0, 1]$ | susceptibility reduction (VE against infection) |
| | $\theta_I \in [0, 1]$ | infectivity reduction (VE against transmission) |
| | $\theta_E \in [0, \infty]$ | relative escape contribution of breakthrough infections |
| epidemiological parameters | $R_0 \in [0, \infty)$ | initial basic reproduction number, here assumed $R_0 > 1$ |
| | $c \in [0, 1]$ | vaccination coverage of the population |
| | $R_e(c) \in [0, \infty)$ | initial effective basic reproduction number, assumed $R_e > 1$ |
| | $\tilde{c} \in [0, 1]$ | vaccination coverage $c$ threshold for $R_e = 1$ |
| escape pressure | $P(t)$ | unscaled and time-dependent, as defined in equation (13) |
| | $\hat{P}_a(c)$ | normalized and cumulative (transient polarised scenario) |
| | $\hat{P}_{a'}(c)$ | normalized and cumulative (transient leaky model of A.2) |
| | $\hat{P}_b(c)$ | normalized and asymptotic (endemic polarised scenario) |
| output parameters | $c_m$ | vaccination level $c$ that maximises the escape pressure |
| | $c_m^\infty$ | vaccination level $c$ that maximises breakthrough cases |
| | $\hat{\theta}_E$ | threshold value of $\theta_E$, defined by $d\hat{P}_*/dc = 0$ at $c = 0$ |
| | $\hat{\theta}_S$ | threshold value of $\theta_S$, defined by $d\hat{P}_*/dc = 0$ at $c = 0$ |
| | $\hat{\theta}_I$ | threshold value of $\theta_I$, defined by $d\hat{P}_*/dc = 0$ at $c = 0$ |

Next, we consider how the escape pressure changes with a variable vaccination coverage *c*, and how this is affected by *θ*_*E*_. We keep *θ*_*S*_ and *θ*_*I*_ fixed for simplicity. Figures 1b, S1b show the escape pressure 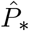 as a function of *c* and *θ*_*E*_, for fixed VEs. Intuitively, the escape pressure decreases to zero as the vaccination coverage approaches 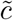, the herd immunity threshold 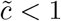 here). When vaccination is slightly below 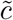, the outbreak is small (a few infections are enough to build herd immunity) so there are fewer opportunities for escape. It is helpful to consider the vaccination coverage that maximises the escape pressure, which we call *c*_*m*_, as it depends on *θ*_*E*_. If *θ*_*E*_ is below a threshold 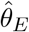, then *c*_*m*_ = 0. The escape pressure simply decreases with *c*, because reducing cases outweighs the escape pressure from breakthrough cases. If 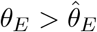, then *c*_*m*_ *>* 0. Breakthrough infections can drive evolution and reducing cases may not suffice to reduce the escape pressure, which has a unimodal shape. We can characterise these results analytically. Here we refer to the endemic scenario (Appendix A.1 discusses the transient scenario). Since *P*_*b*_(*c*) is concave (*d*^2^*P*_*b*_*/dc*^2^ < 0), by (17), the condition for a peak of the escape pressure at a non-zero vaccination level (*c*_*m*_ *>* 0) is

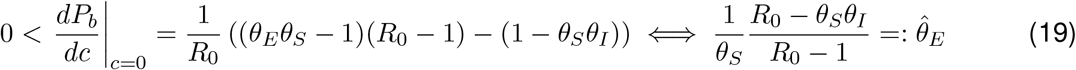

Figure S5 plots 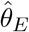, as given by (19). Interpreting 1 − *θ*_*S*_*θ*_*I*_ as the transmission-blocking and *θ*_*E*_*θ*_*S*_ 1 as the escape boost provided by the vaccines, the condition (19) can be rewritten as (escape boost)*×*(*R*_0_ − 1) *>*(transmission-blocking). If (19) is satisfied, we solve *dP*_*b*_*/dc* = 0 folr the maximiser of the escape pressure 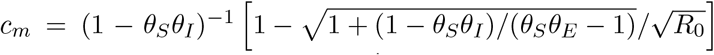. As expected, *c*_*m*_ continuously increases with *θ*_*E*_ from zero (at 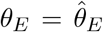). In the limit *θ*_*E*_ *→ ∞*only breakthrough infections contribute to *P*_*b*_, the analytical expression for *c*_*m*_ tends to 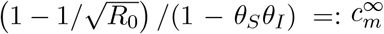. This number 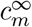 can be obtained directly as the vaccination coverage that maximises the total number of infections in vaccinated hosts. If herd-immunity from vaccination alone is not possible 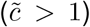, Figure S6 shows that a fully vaccinated population (*c* =1) can have the largest escape pressure. However, this does not happen 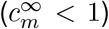 if and only if 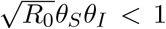, which holds if the threshold coverage for *R*_*e*_ < 1 is less than one 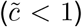, because then *R*_0_*θ*_*S*_*θ*_*I*_ < 1 < *R*_0_. Therefore, unsurprisingly, the escape pressure cannot peak at *c* = 1 if 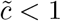 (since *c* = 1 prevents the epidemic).

Finally, we study how each of *θ*_*S*_, *θ*_*I*_ affects the escape pressure as the vaccination coverage varies. When infections in the vaccinated drive selection (*θ*_*E*_ is large), *θ*_*S*_ contributes more to the escape pressure than *θ*_*I*_. Both reduce the total number of cases at the same rate, through *R*_*e*_ = *R*_0_(1 − *c*(1 −*θ*_*S*_*θ*_*I*_)). Additionally, *θ*_*S*_ (partially) protects the vaccinated from infections, so it lowers the proportion of the total cases in the vaccinated. Hence, if *θ*_*E*_ *>* 1, a vaccine that reduces susceptibility has a quantitatively stronger effect on the escape pressure than a vaccine that reduces infectivity. Figures 2, S2, S3, S4 demonstrate that the escape pressure is more sensitive to *θ*_*S*_ than *θ*_*I*_. In particular, Figures 2, S2 show that a sufficiently low *θ*_*S*_ can lead to the escape pressure peaking at no vaccination (*c*_*m*_ = 0), while for the same background parameter values the escape pressure peaks at intermediate vaccination levels (*c*_*m*_ *>* 0), regardless of *θ*_*I*_. We can define thresholds values 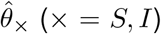 similar to 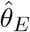 (see Figure 1b, S5), such that 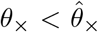 is the condition for the escape pressure to peak at no vaccination (*c*_*m*_ = 0). Figures 2a, S2a, S3a all have 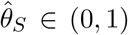, while 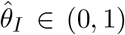 only appears in Figure S3b. In the endemic scenario, (19) gives 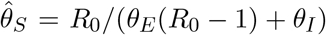 and 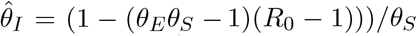. The expression for 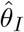 can be negative, but *θ*_*I*_ is always positive. Therefore, if 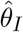 is negative, the escape pressure peaks at intermediate levels, as in Figure 2b. On the contrary, 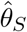 is always positive, so if vaccines fully prevent infection (*θ*_*S*_ = 0), vaccination always reduces the escape pressure (*c*_*m*_ = 0).

**Figure 2:**
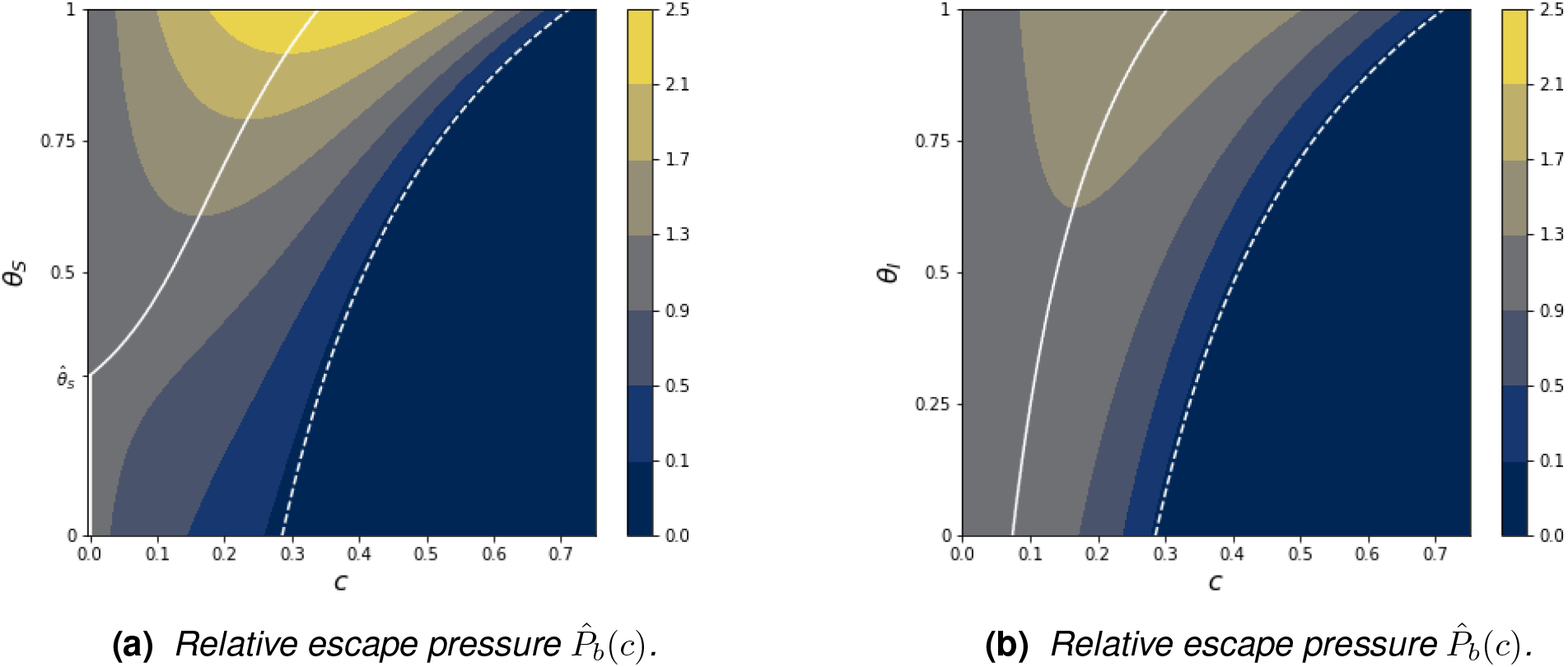
Relative escape pressure 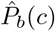 at fixed R_0_ = 1.4 and a high relative contribution to escape from the vaccinated, θ_E_ = 10. (a) has fixed θ_I_ = 0.6 and the susceptibility VE θ_S_ varies. (b) has fixed θ_I_ = 0.6 and the infectiousness VE θ_I_ varies. On both (a) and (b), the white solid curves mark the vaccination coverage, c_m_, at which the escape pressure peaks; the dashed lines give the herd-immunity threshold. Considered together, (a) and (b) show that there is more variation in the relative escape pressure if θ_S_ varies and θ_I_ is fixed.

## 4 Discussion

### 4.1 Summary of findings

Introducing a new parameter, *θ*_*E*_, we have explored the effect of the contribution to immune escape of cases in vaccinated hosts, relative to the unvaccinated. Taking a parsimonious modelling approach we have studied how the population escape pressure depends on the vaccination coverage, the vaccine efficacies and *θ*_*E*_. We have found that *θ*_*E*_ is critical to determine how the escape pressure depends on the vaccination coverage. If *θ*_*E*_ is low, escape decreases with vaccination. As *θ*_*E*_ increases, so that vaccinated hosts contribute relatively more to escape, escape is largest at intermediate vaccination levels. Therefore, our results show that models of immune escape should take into account the relative contribution to escape of cases in hosts with imperfect immunity. Our results also show mathematically that vaccines which are effective in reducing transmission can generally reduce the population escape pressure, even if they favour escape strains in individual hosts.

### 4.2 Implications for modelling evolution with imperfect immunity

Our results highlight not only the importance of *θ*_*E*_ itself, but also the role in reducing escape of *θ*_*S*_, the susceptibility vaccine efficacy (VE), especially when vaccinated hosts contribute relatively more to escape (*θ*_*E*_ *>* 1). Our results show mathematically the need to consider separately the VEs against infection (susceptibility) and onwards transmission (infectiousness). A purely epidemic model which does not consider immune-induced escape only needs the overall transmission-blocking VE [4]. However, our escape pressure is more sensitive to a reduction in susceptibility than infectivity. The suceptibility VE not only reduces infections but specifically protects the vaccinated, who may exert more selection pressure. Traditional models assume that partial immunity confers only reduced susceptibility or infectivity [16]. Often the dynamics are qualitatively the same [25], so the choice of VE is usually based on tractability [16]. Our work suggests that evolutionary models should split infections by immune status, so that *θ*_*E*_ can be used, and consider the susceptibility VE separately from the transmisibility VE, so that *θ*_*S*_ and *θ*_*E*_ can regulate together the escape pressure. These points may also apply in multi-strain models where partial cross-immunity drives evolution [16]. Furthermore, the contribution to escape of an infected host may change with different vaccine types or number of doses, so along different VEs, different *θ*_*E*_s may be needed. Parameters akin to *θ*_*E*_ could also be appropriate for other phylodynamic models.

### 4.3 Relation to existing results

As in previous work [8], [3], in our models intermediate vaccination levels could lead to the largest selection pressure for antigenic escape. The same conclusion holds for a wide range of *θ*_*S*_ and *θ*_*I*_ values [4] when dividing the population in two subgroups with different contact rates. This population-level result is analogous [4] to the within-host phylodynamic trade-off between viral load and selection pressure [23]. Our works shows that a population-level trade-off can appear without mixing heterogeneity, stochasticity, or detailed within-host dynamics. Past work implicitly uses fixed choices of *θ*_*E*_ or the VEs (or both). We instead specify the region of (*θ*_*E*_, *θ*_*S*_, *θ*_*I*_) parameter space in which intermediate vaccination levels maximise the escape pressure. Beyond this region, vaccination reduces the escape pressure, as in [15].

### 4.4 Limitations and further work

Due to the simplicity of our approach, many epidemiological and evolutionary processes could modify our results. We assume that there are no changing mitigation measures, behavioural changes or seasonality effects. Vaccination comes through a single dose of a unique vaccine, administered before the outbreak or at birth. Further work could consider ongoing vaccination, as in [8]. We also neglect the waning of vaccine immunity and assume complete immunity after infection. Infections in individuals with partial or waned immunity may contribute with a different weight to the escape pressure [5]. Heterogeneity in mixing, susceptibility and transmissibility could all play a role in evolution. It may be best to vaccinate first those with more contacts [4], or immunocompromised hosts, who might contribute significantly more to the escape pressure [26]. Moreover, unlike in COVID-19 [22], both scenarios explained here assume that vaccination does not change the recovery rate and that the disease is not fatal. Disease-induced mortality, different infectious periods and hospitalisations would break the proportionality (4), since these effects would be larger in the unvaccinated. Similarly, lifting the well-mixing assumption and including any age or spatial structure could change these patterns. These assumptions are a tradeoff in favour of tractability and simplicity: the main objective of our of work is to gain as much insight as possible about the qualitative effect of *θ*_*E*_ on the escape pressure.

Our escape pressure (13) is just a minimal abstraction of the interplay between vaccination and vaccine escape that might occur in real-life. Our work ignores the invasion dynamics of escape strains [6], but simply focuses on the pressure to generate one. An escape strain might be unable to grow if few individuals remain susceptible. In the transient scenario, this could occur if a strains appears late in the epidemic; in the endemic scenario, if the invasion fitness of the escape strain is not large enough for it to replace or coexist with the resident strain. Similarly, stochasticity could drive escape strains extinct shortly after their generation. For example, [8] finds that escape strains appearing in lockdowns are unlikely to survive stochastic extinction. Simple approaches to consider the outcome of the new variant [8], [6] could be incorporated in our models. However, this is a complex process [3], so a full description will require significant further development of models.

## Data Availability

There is no data used in this manuscript. The results are reproducible only with the information presented in the manuscript.

## Acknowledgements

MAG is supported by the Gates Cambridge Trust (grant OPP1144 from the Bill & Melinda Gates Foundation).

## Declaration of interests

We declare we have no competing interests.

## A Mathematical proofs

### A.1 Polarised vaccine immunity

Using the notation of the main text, define column vectors **u**(*t*) and **v**(*t*) as

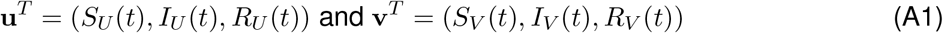

where **x**^*T*^ denotes the transpose of a vector **x**. Consider an ODE system

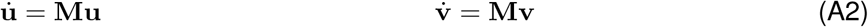

for a time-dependent matrix **M**, which possibly depends on **v** and **v**. Suppose initial conditions

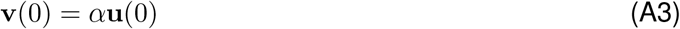

for some constant *α*. **û** (*t*) := *α*^−1^**v**(*t*) and **u**(*t*) obey the same first order differential equation (A2) and initial condition **u**(0) = **û** (0). Hence (by uniqueness of solutions), **û** (*t*) = **u**(*t*). Thus **v**(*t*) = *α***u**(*t*).

### Analytic results for the transient polarised model

The system (1)-(3) can be written in the form (A1)-(A3) with 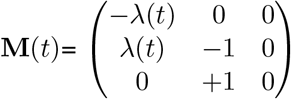 and *α* = *cθ*_*S*_*/*(1 − *c*). Thus (*S*_*V*_ (*t*), *I*_*V*_ (*t*), *R*_*V*_ (*t*)) = *cθ*_*S*_(1 − *c*)^−1^(*S*_*U*_(*t*), *I*_*U*_(*t*), *R*_*U*_(*t*)). This relation (4) allows us to derive (5), which yields

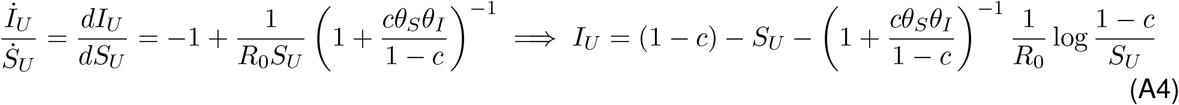

where the initial conditions (6) set the integration constant. The final size of the susceptible population, 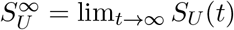, is given implicitly by taking *t → ∞* and setting 0 = lim_*t→∞*_ *I*_*U*_(*t*):

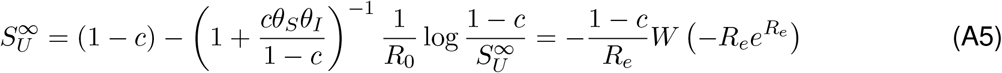

The second equality expresses 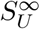 in terms of the Lambert-W function [21] and *R*_*e*_. From 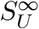 we obtain (14) for the escape pressure *P*_*a*_(*c*), which we study here. For simplicity, we set *W ≡ W* (−*Re*^−*R*^) and *R ≡ R*_*e*_.

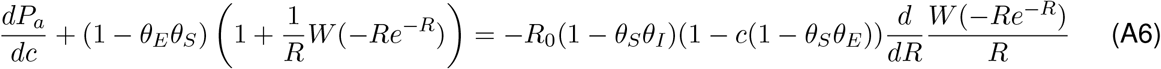

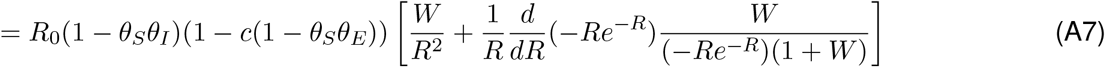

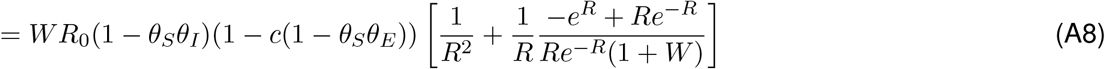

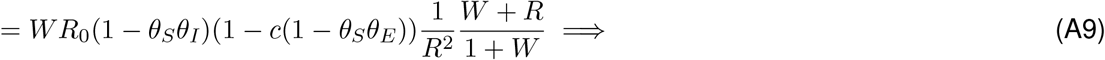

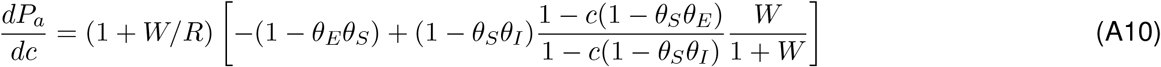

See, e.g., [21] for the derivative of the Lambert-W function used in (A7). We prove that there is either none or one root (*c* = *c*_*m*_) solving *dP*_*a*_*/dc* = 0 in 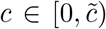. *W* ≥ −1, and hence (1 + *W/R*) *>* 0, because *R >* 1 for *c* < *c*_*m*_. Thus it suffices to consider the sign and monotonicity of the bracketed (right-most) term in (A10). First suppose (1 −*θ*_*S*_*θ*_*E*_) *>* 0, which means (1 − *c*(1 −*θ*_*S*_*θ*_*E*_)) *>* 0. Since *W/*(*W* + 1) < 0, *dP*_*a*_*/dc* < 0. So if *θ*_*E*_ < 1*/θ*_*S*_, vaccination reduces the escape pressure regardless of the coverage *c*. Now consider *θ*_*E*_*θ*_*S*_ *>* 1. *R* decreases with *c* and *R >* 1 for 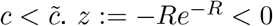 increases with *R >* 1, so *z* decreases with 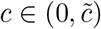. The Lambert W function *W*(*z*) increases with its argument *z ∈* (−1*/e*, 0), so it decreases with *c. W/*(1 + *W*) is an increasing function of *W* ∈ (0, −1) and hence it is a negative, decreasing function of *c*. These imply

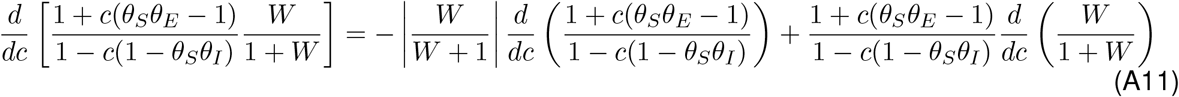

where the first derivative in the RHS is positive and the second is negative. Hence the LHS is negative. Thus *dP*_*a*_*/dc* is a decreasing function of *c*, with at most one root, *c*_*m*_. The if and only if condition for *c*_*m*_ *>* 0 is *dP*_*a*_*/dc >* 0 at *c* = 0. Evaluating (A10) at *c* = 0, the condition for *c*_*m*_ *>* 0 becomes

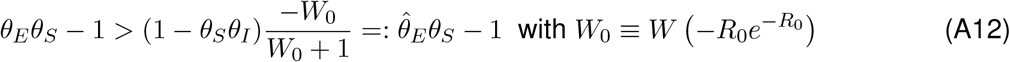

(A12), as (19), can be written in the form (escape boost) *> g*(*R*_0_)(transmission blocking) for a function *g*. Here *g*(*R*_0_) = −*W*_0_*/*(1 + *W*_0_); while in the endemic scenario, *g*(*R*_0_) = 1*/*(*R*_0_ − 1).

### Analytic results the endemic models

The SIRS system (7)-(9) can be written in the form (A1)-(A3) with 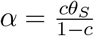 and 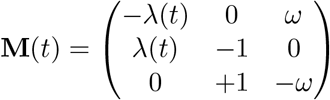.

Thus, (4) holds. We write the system with natural dynamics (10)-(12) as 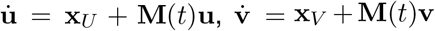, with 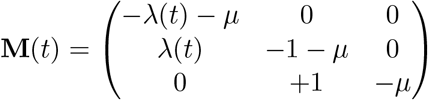 and 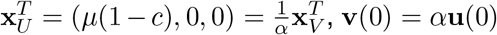 for *α* = *cθ*_*S*_/(1 − *c*). Both **u**(*t*), **û** (*t*) := *α*^−1^**v**(*t*) solve the initial value problem 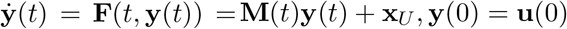. By uniqueness of solutions, **u**(*t*) = **û** (*t*), and hence (4) still holds.

### A.2 Analytical results for leaky vaccine immunity

We modify the SIR model of Section 2.1 to have “leaky” immunity, instead of polarised [16]:

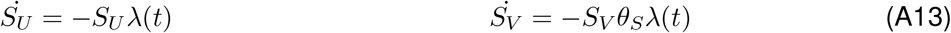

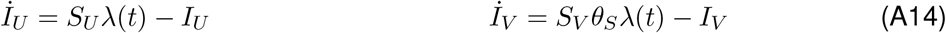

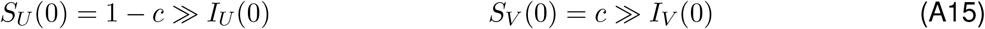

For *c* = 1 or *c* = 0, (A13)-(A15) become a standard SIR model: 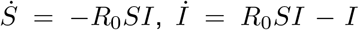, *S*(0) = 1 ≫ *I*(0) *>* 0 if *c* = 0, and the same but with *R*_0_ *→ R*_0_*θ*_*S*_*θ*_*I*_ for *c* = 1. We assume *R*_0_ *θ*_*S*_*θ*_*I*_, *R*_0_ *>* 1 so that the outbreak grows in both situations. The final sizes are

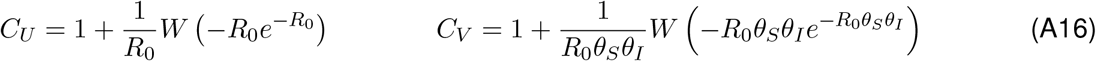

respectively, where *W* is the Lambert-W function [21]. From (A16) and (18) we find that the relative escape pressure in a fully-vaccinated population under the “leaky” immunity assumption is 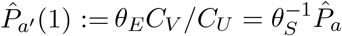, proportional to the relative escape pressure under polarised immunity. Figures S1a, S1c are hence identical, except for the vertical variables, *θ*_*E*_ and *θ*_*S*_*θ*_*E*_.

For *c ∈* (0, 1), from (A13), 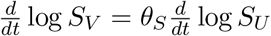 and so 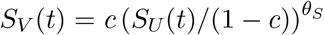, which deviates from (4). However, we can still study the system through *S*_*U*_(*t*) and *λ*(*t*) = *R*_0_(*I*_*S*_ + *θ*_*I*_*I*_*V*_):

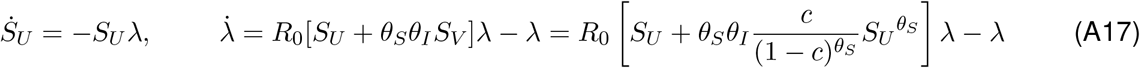

Using the chain rule and initial conditions (A15), 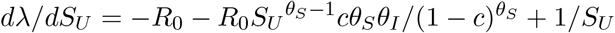, so 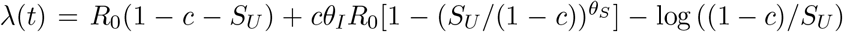. Setting lim_*t→∞*_ *λ*(*t*) = 0, (A18) defines 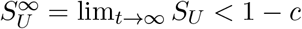, the final size of the unvaccinated susceptible compartment:

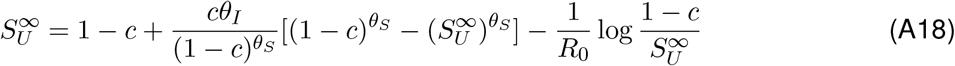

The cumulative number of infections in each group are 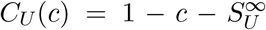 and 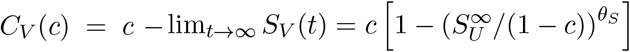 so, for 0 < *c* < 1,

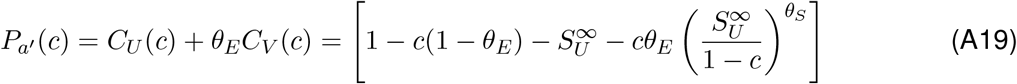

and where 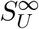 is the unique solution in (0, 1 − *c*) of (A18) for *R*_*e*_ *>* 1. For *R*_*e*_ ≤ 1, we set *P*_*a*_*′* = 0. Figures S1b, S1d are the equivalent of Figure 1b for the “leaky” and polarised transient models. The effect of *θ*_*E*_ is qualitatively the same in all models, regardless of vaccination coverage.

## S Supplementary Figures

**Figure S1:**
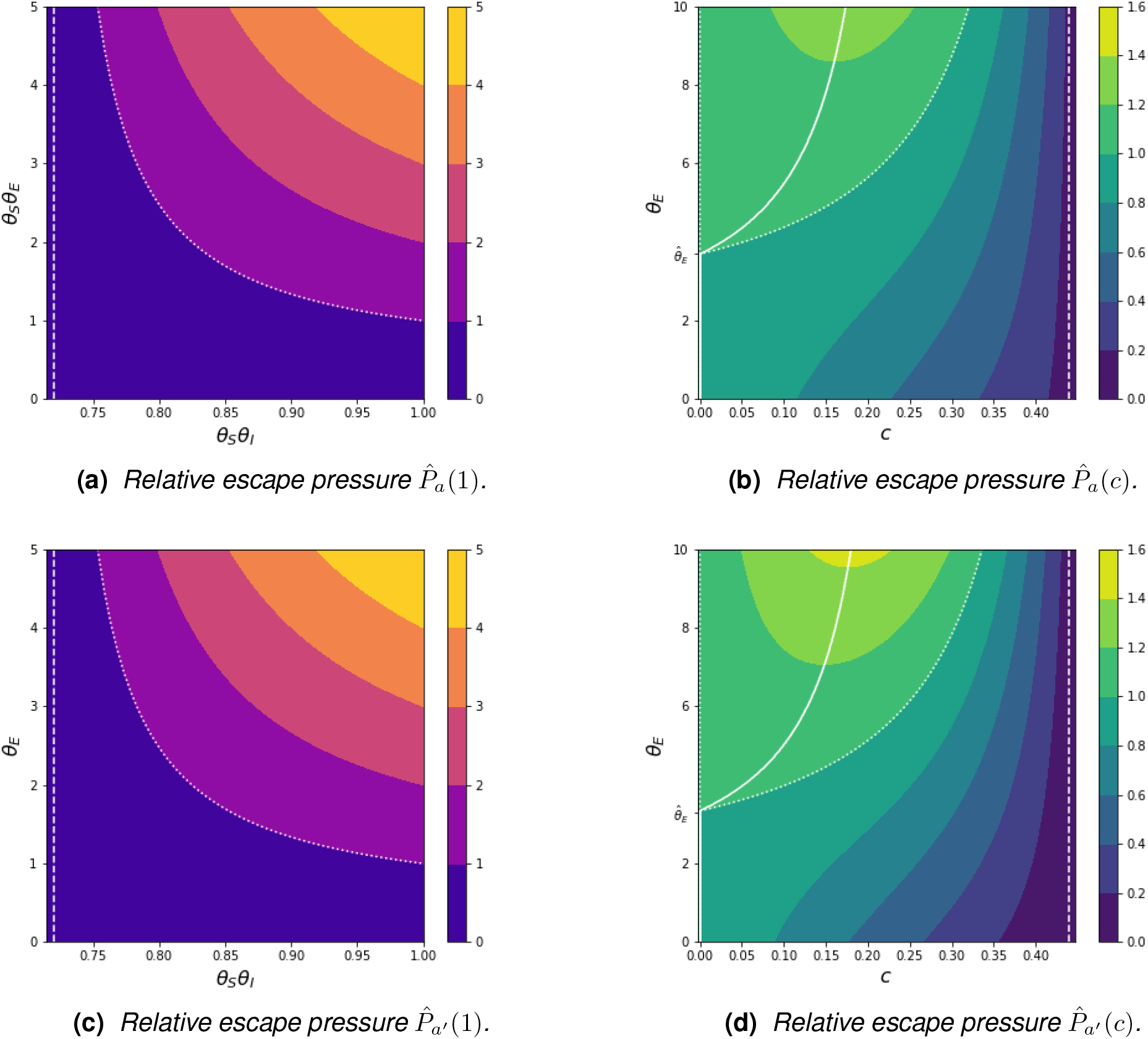
As Figure 1 but for the transient scenario. The qualitative behaviour is the same as in the endemic scenario. Top row is with polarised immunity, bottom row is for leaky immunity. The escape pressure is larger with leaky immunity instead of polarised immunity (as seen in the different variables in the vertical axis of (c) or the maximum values of 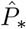 attained in (d)).

**Figure S2:**
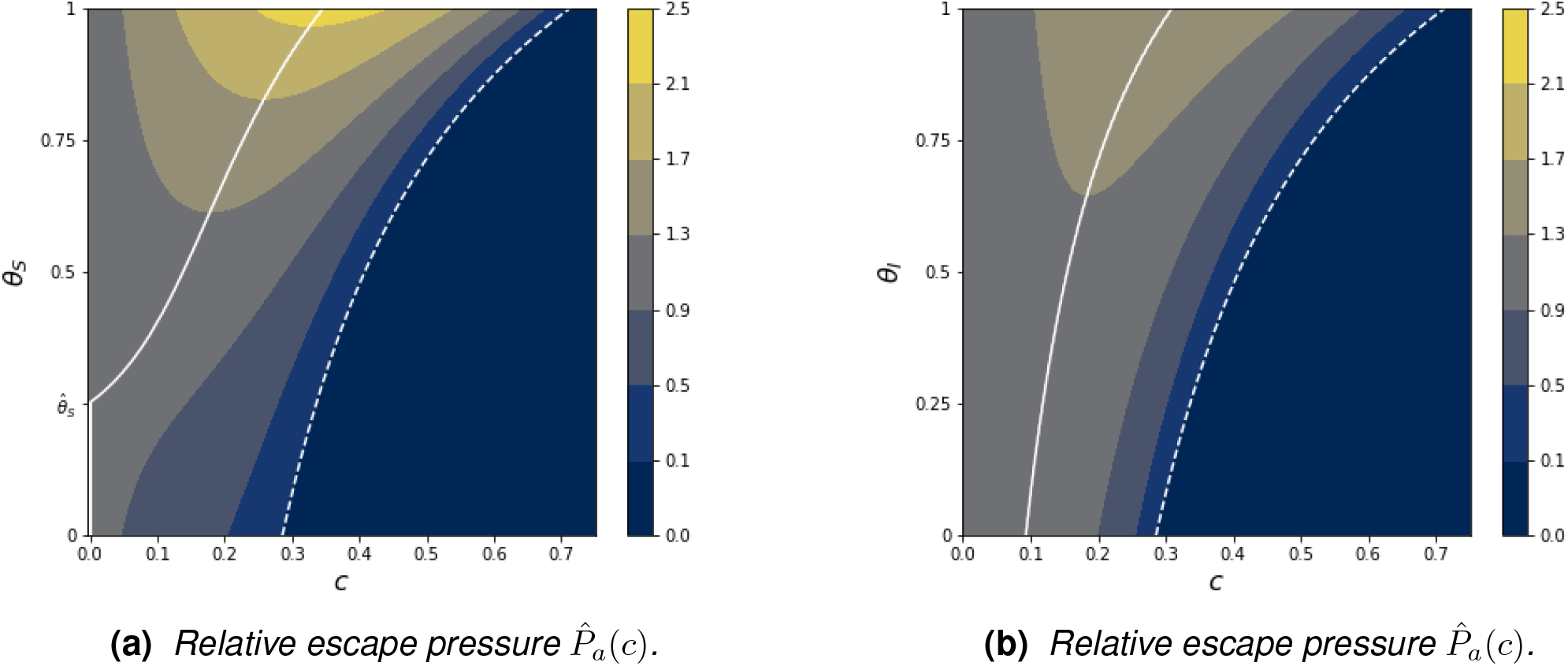
As Figure 2, but for the transient scenario with polarised immunity. The qualitative behaviour is the same as in the endemic scenario. The same behaviour appears in the transient scenario with leaky immunity (plots not shown).

**Figure S3:**
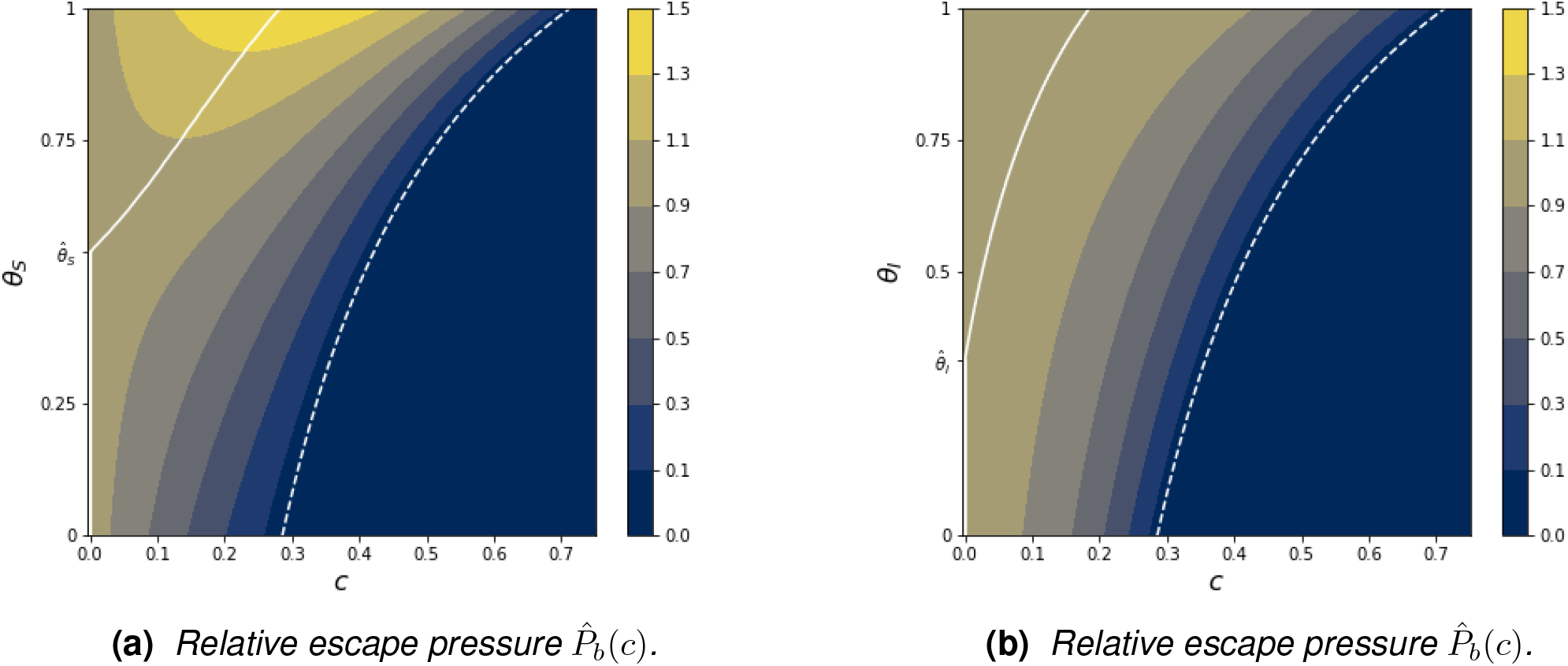
As Figure 2, but with a lower relative contribution to escape from vaccinated hosts: θ_E_ = 5 instead of θ_E_ = 10. Here vaccination that fully blocks infection (θ_S_ = 0) or onward transmission (θ_I_ = 0) reduces the escape pressure at any vaccination coverage (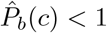 for *c* > 0). In other words, here 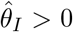. However, the escape pressure is still more sensitive to θ_S_ than θ_I_. The same behaviour appears in the transient scenario with leaky or polarised immunity (plots not shown).

**Figure S4:**
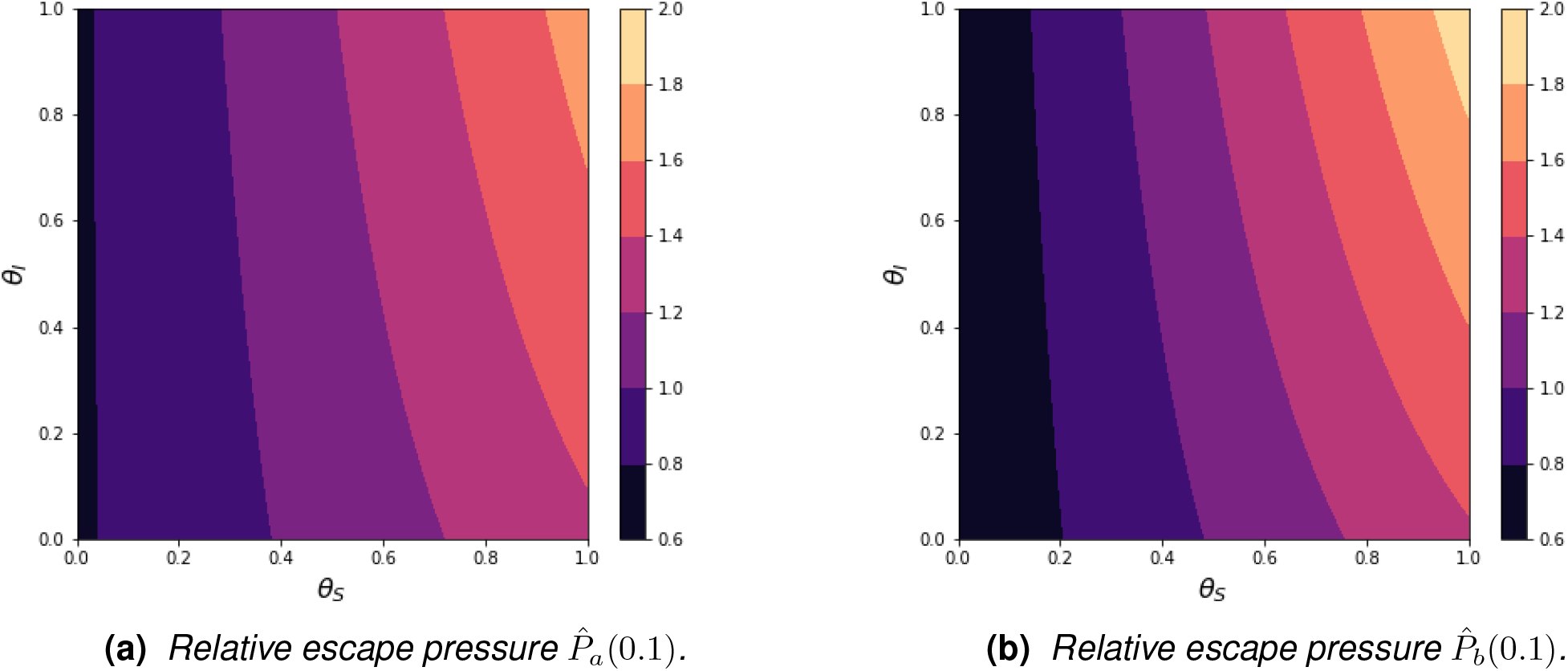
Relative escape pressure 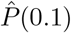 as a function of the vaccine efficacies θ_S_ and θ_I_ for a fixed vaccination coverage c = 0.1. (a) is for the transient scenario and (b) is for the endemic scenario: the qualitative results are the same. As expected, reducing θ_S_ or θ_I_ decreases the escape pressure, because this corresponds to more effective vaccines. However, the contour lines that gives constant escape pressures are roughly vertical, meaning that the escape pressure decreases is much more sensitive to θ_S_ than θ_I_. As in Figures 2, S2, R_0_ = 1.4 and θ_E_ = 10. The same behaviour appears in the transient scenario with leaky immunity (plots not shown).

**Figure S5:**
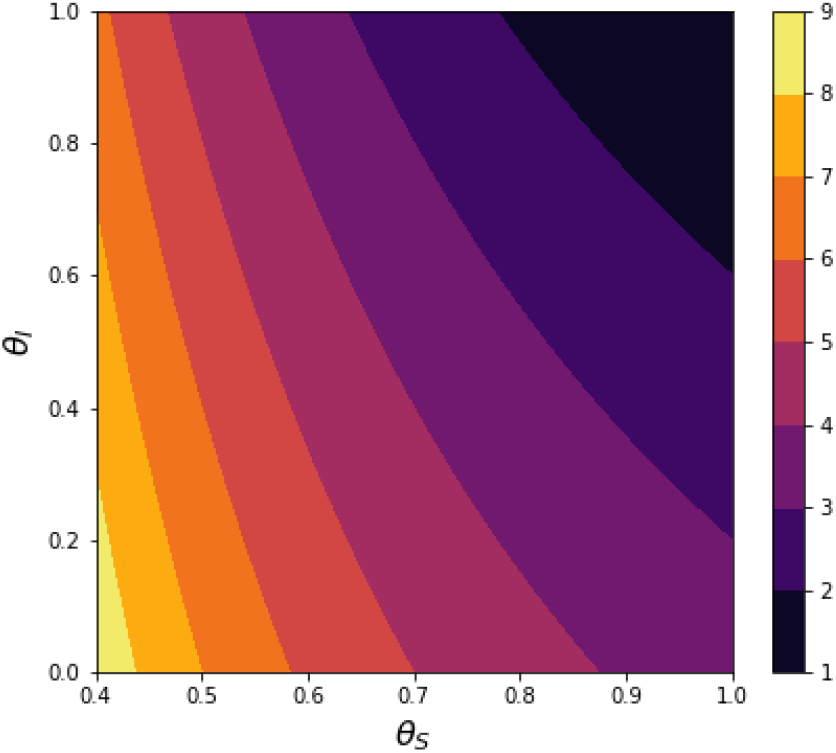
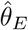 (the value of the relative contribution to escape from the vaccinated above which intermediate coverages maximise the escape pressure) in the endemic scenario, as given by (19). We only plot values of the susceptibility VE θ_S_ above 0.4, because 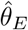 quickly grows without bounds as θ_S_ → 0 (the vaccines become better at preventing infection). In other words, 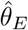 is more sensitive to θ_S_ than θ_I_, as 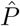 (see, e.g., Figure S4). R_0_ = 1.4.

**Figure S6:**
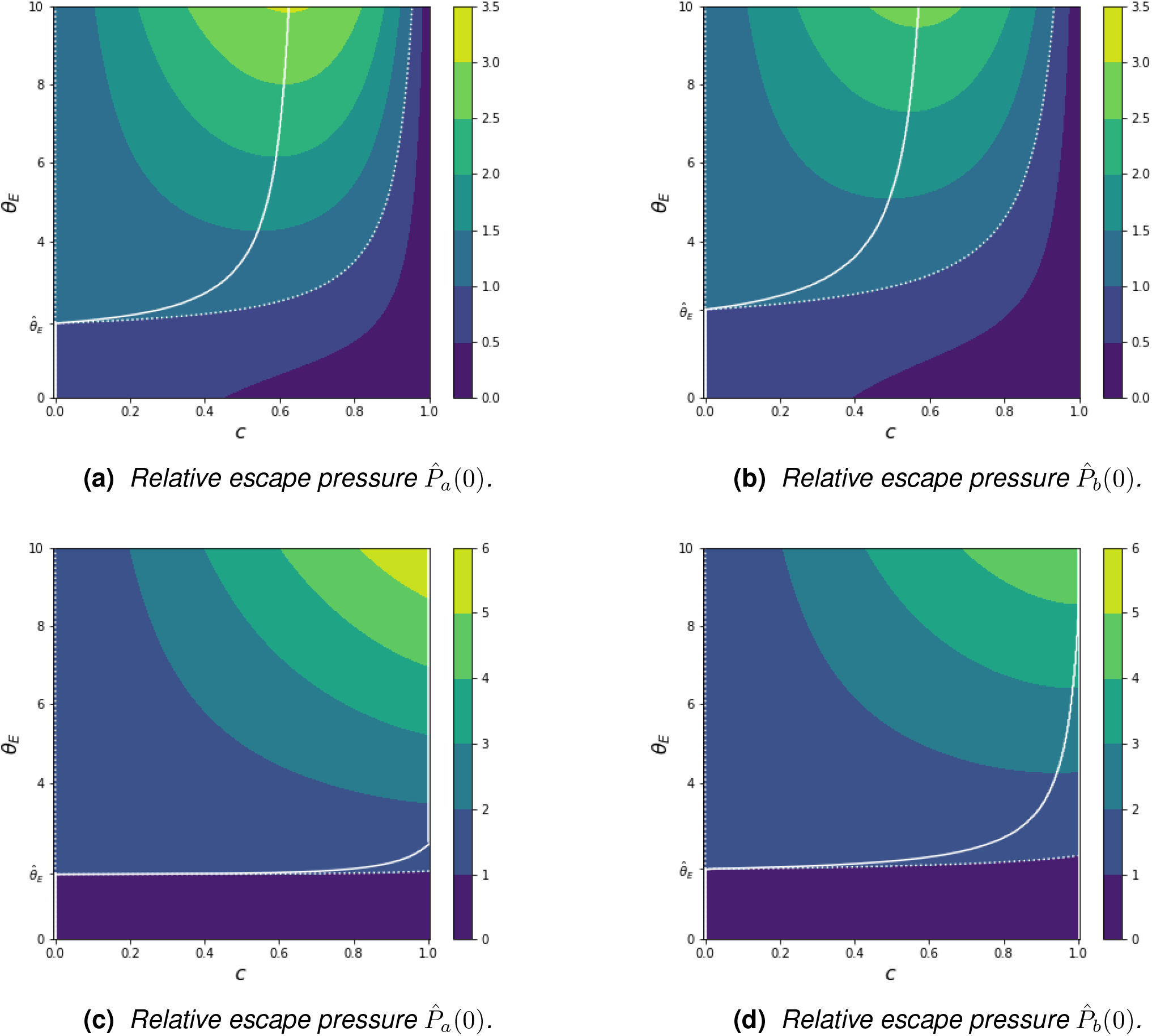
As Figures 1b, S1b, but with different R_0_ such that vaccination alone cannot achieve herd-immunity 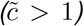. (a), (b): R_0_ = 2.8. The escape pressure is non-zero even in a fully vaccinated population (c = 1), because there is still transmission. Nevertheless, the vaccination coverage that maximises the escape pressure is always less than 1, so the escape pressure does not peak at a fully vaccinated population regardless of how much cases in vaccinated individuals contribute to the escape pressure (c_m_ < 1), 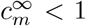 is the vertical asymptote of the white curve, the limit of c_m_ as θ_E_→ ∞. (c), (d): R_0_ = 9. For large θ_E_, the escape pressure increases monotonically with the vaccination coverage c and is largest at a fully-vaccinated population (c = 1). As with the other plots of this paper, the qualitative behaviour of the transient (left column) and endemic (right column) scenarios is the same. We do not show plots for the transient scenario with leaky immunity, but the qualitative behaviour is the same.

## Notes

### Competing Interest Statement

The authors have declared no competing interest.

### Funding Statement

This study did not receive any funding.

### Summary of Updates

Restructured Methods section, now including a paragraph discussing the escape pressure as a function of time. Clarified escape pressure used in transient and endemic scenarios. Other minor edits for clarity.

